# Three pictures of COVID-19 behavior in Italy: similar growth and different degrowth

**DOI:** 10.1101/2020.05.09.20096149

**Authors:** A. Feoli, A. L. Iannella, E. Benedetto

## Abstract

In this short note, we present an analysis of the data regarding Coronavirus (COVID-19) in Italy. We have used the official database provided by the Italian Civil Protection. Since the total number of people infected by the virus is uncertain, we have considered the trend of ICU patients, ICU patients plus deaths, and finally the sum of hospitalized patients plus the deceased. The growth of the corresponding curves is similar for all the three graphs while the trend after the turning point is completely different. We find that the curve of ICU patients can be a useful tool to monitor the behavior of epidemic and a model to predict the future evolution of COVID-19.

## 1 Introduction

In late January there were the first two cases of COVID-19 in Italy, when two tourists tested positive for the virus. Not many days later, an Italian man was hospitalized and confirmed as a third case. Unfortunately, the virus began to spread and in early March all Italian regions had been infected. Moreover, ICU (Intensive Care Unit) patients began to increase and on February 22nd there was the first official case of death caused by the virus. This series of dramatic events has forced the Italian Government to take, from the month of March, a series of restrictive measures. In other words, a national quarantine was imposed limiting the possibility of people movements, except in cases of extreme necessity. The first lockdown initially concerned some “red zones” in the northern part of Italy, but it was gradually extended to the entire country. We have analyzed the official data provided by the Italian Civil Protection starting from March 2nd to May 1st [1]. The data of Deaths occurred on May 2nd are not correct because, following the interviews released by members of Italian task force, 282 deaths registered for May 2nd are, on the contrary, relative to the last days of April but they have not specified the details of this error, so it is possible that the data of deaths for April 30th and May 1st are slightly underestimated (for the same reason the number of total deaths registered in the table of March 26th were corrected from 8165 to 8215). Anyway, we have decided to interrupt our statistical analysis also considering the fact that for May 4th the beginning of a new phase is fixed, with less restrictive rules for people movements. The change of boundary conditions, obviously, will cause a correspondent change in the statistical curves.

**Table 1.** Parameters of Gaussian functions for the first 28 points of the Epidemic

| $y(x) = Ae^{\frac{-(x-B)^2}{C^2}}$ | ICU Patients | ICU Patients + Deaths | H Patients + Deaths |
| --- | --- | --- | --- |
| $A$ | $3900 \pm 21$ | $18507 \pm 334$ | $45697 \pm 727$ |
| $B$ | $30.29 \pm 0.18$ | $37.36 \pm 0.38$ | $34.00 \pm 0.41$ |
| $C$ | $16.60 \pm 0.15$ | $17.26 \pm 0.21$ | $17.09 \pm 0.27$ |

Note: ICU Patients are Patients in Intensive Care Units, while H Patients are composed by Hospitalized Patients with symptoms and Patients in Intensive Care Units.

Mathematical analysis of experimental data concerning COVID-19 has recently become of great interest [2-5]. The numbers regarding people who tested positive for the virus are certainly unrealistic. There are many asymptomatic but infected people that are not counted by the official data, because they have not been discovered as positive to a test. The amount of this hidden cases is unknown. It is generally overestimated when the analysis regards the percentage of deaths compared to the infected. Actually, on May 2nd there were in Italy 209328 total cases with 28710 deaths. So, the mortality rate is 13.7% and it is too high, but considering a great number of hidden infected people, the mortality rate decreases and becomes in agreement with the one of other countries. On the contrary the infected that were not detected are underestimated when the analysis is done to show that their number is too low to achieve herd immunity in a short time. For this reason we have analyzed the data that seem more solid like the hospitalized patients (hospitalized patients with symptoms and patients in ICU) and the deceased, even if, especially in some areas of Lombardia, the number of deaths is clearly underestimated. In particular, the great numbers of ICU patients and deceased are important data also because they represent the peculiarity of COVID-19 with respect to the evolution of a virus of the annual well known flu.

We have followed the time evolution of three curves. The trend of ICU patients (*curve a*), ICU patients plus deaths (*curve b*), and finally the sum of hospitalized patients and the deceased (*curve c*). In Sect.2 we present three steps of evolution of these curves and in Sect.3 we discuss the obtained results, suggesting which curve can be used to monitor the evolution of COVID-19.

## 2 Three pictures in three steps

We report the behavior of the virus expansion in Italy during two months from March 2nd to May 1st. In principle, the curves could be used not only as thermometers of the virulence of COVID-19 and its impact on Italian population, but also to predict the future development of epidemic. Furthermore what we learn from the study of these curves can be useful for other countries, where the epidemic has begun later, or for a possible new increase of cases in Autumn.

In the first step we note that all the three curves show a Gaussian behavior that is very impressive for the coincidence of the experimental points with the best fit curves that are calculated using the routine “NonlinearRegression” of MATHEMATICA and refer to the first 28 experimental points. Given the surprising overlap of the curve on the experimental data, in this first phase, all the three best fit line can be used to make short term (two or three days) forecasts of the behavior of the epidemic. The values of the three parameters of the best fit function are reported in **Table 1**. Regarding some long term forecasts, from the analysis of the first step (**Fig. 1**), the extrapolation from the experimental data allows to predict a peak for the *curve a* for day 30 (i.e. March 30th), for the *curve b* for day 37 (i.e. April 6th) and for the *curve c* for day 34 (i.e. April 3rd). This prediction appears reasonable and in agreement with what we expected. Of course, the *curve b* and *curve c*, after the peak, cannot decrease until the point zero but, at the most, their degrowth will stop at the total number of deaths.

**Figure 1.**
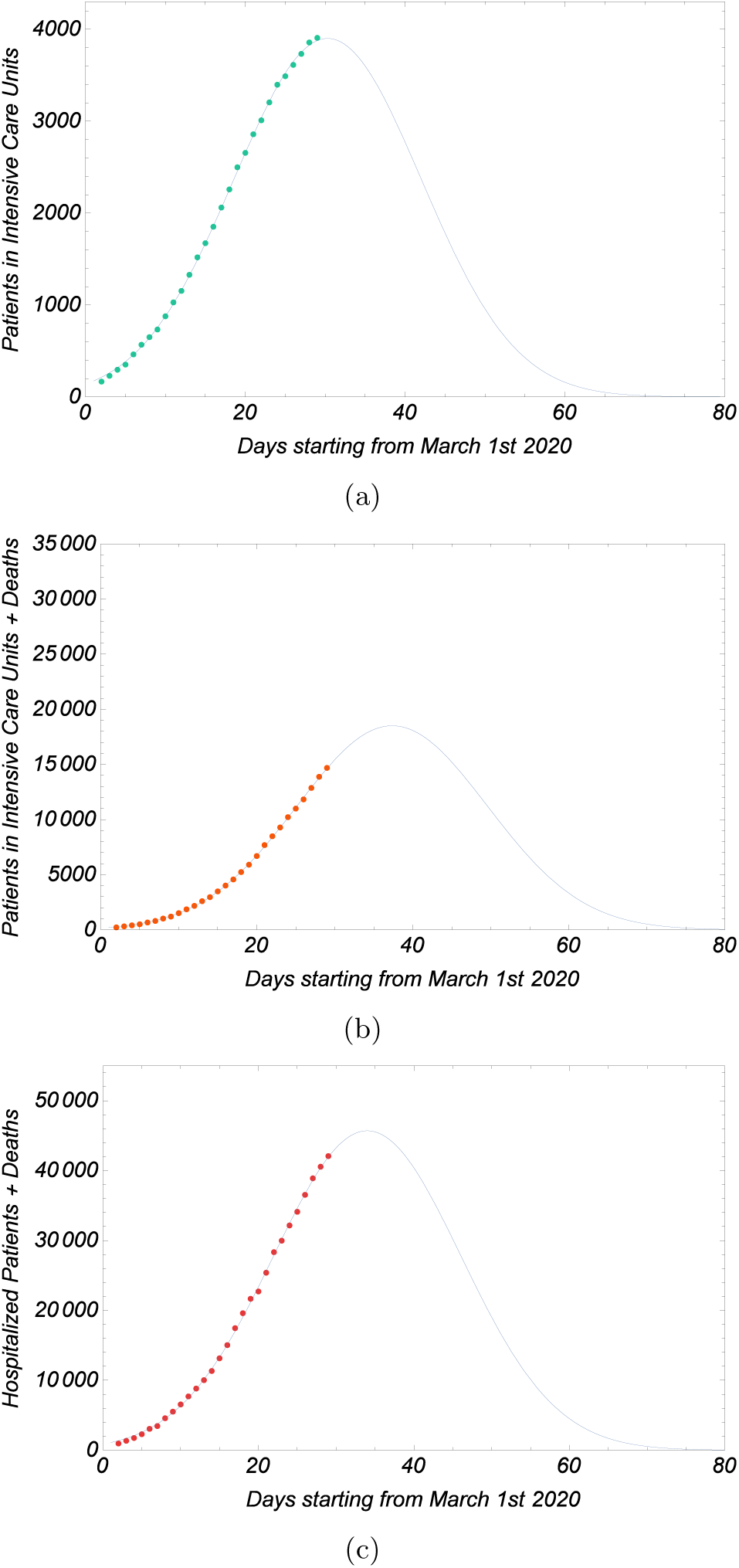
The Epidemic Spreading referred to the first 28 experimental points: (a) Patients in Intensive Care Units, (b) Patients in Intensive Care Units and Deaths, (c) Hospitalized Patients (Hospitalized Patients with symptoms and Patients in Intensive Care Units) and Deaths.

The second step shows the same curves after other eight days (**Fig. 2**), two days after April 3rd, when the *curve a* has experimentally achieved the peak of the Gaussian. Hence for the *curve a* the prediction was wrong of only four days, but the decrease of the curve has really begun. On the contrary, for *curves b* and *c* the growth goes on and the theoretical peak has moved forward and it is on day 40 for *curve b* and day 36 for *curve c*. The best fit values of the parameters are listed in **Table 2** and we note also that the maximum of the *curve a* in **Table 1** (3900 ± 21) predicts the experimental peak (4068) with an acceptable error of 3.6%.

**Figure 2.**
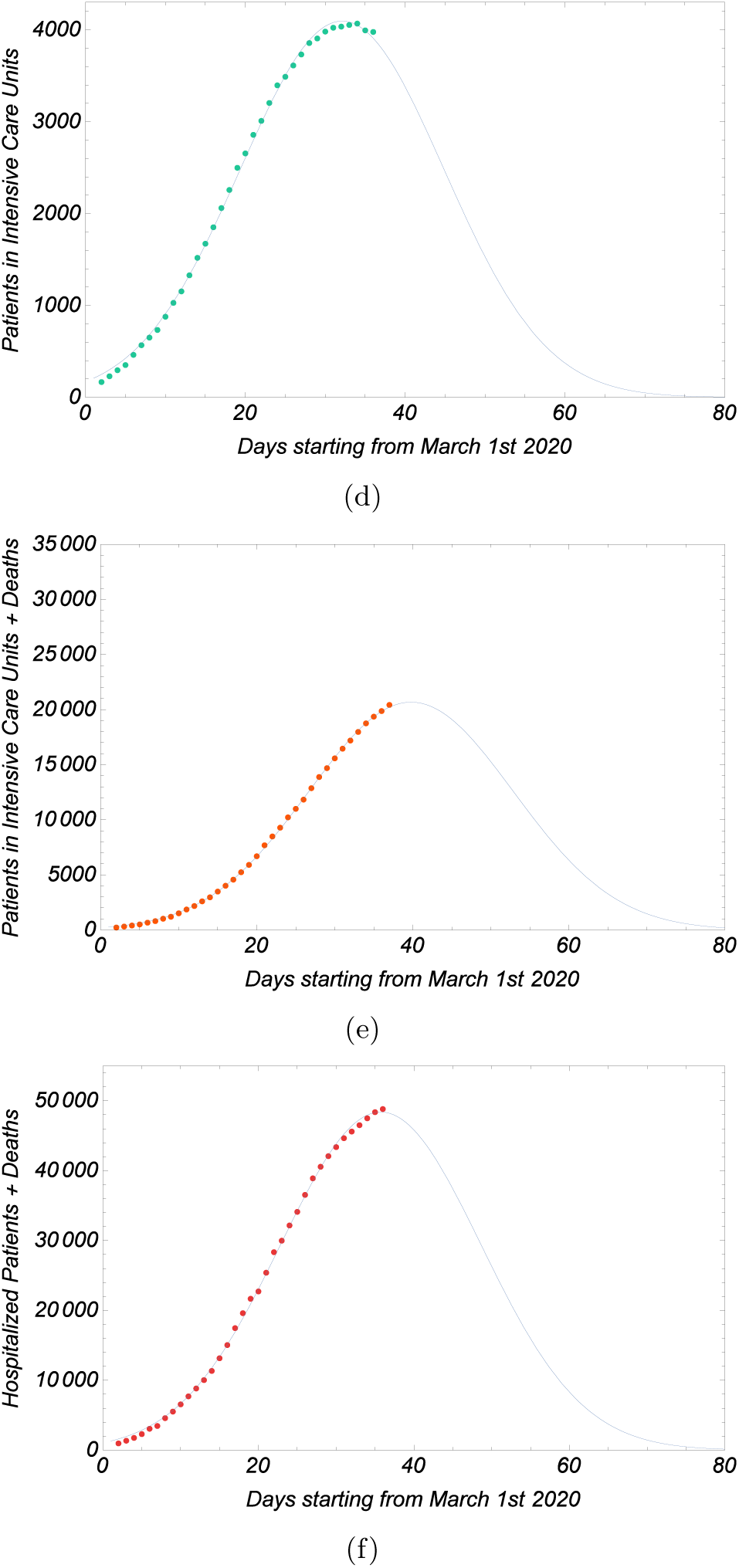
The Epidemic Spreading referred to 36 experimental points: (a) Patients in Intensive Care Units, (b) Patients in Intensive Care Units and Deaths, (c) Hospitalized Patients (Hospitalized Patients with symptoms and Patients in Intensive Care Units) and Deaths.

**Table 2.** Parameters of Gaussian functions for 36 points of the Epidemic Spreading.

| $y(x) = Ae^{\frac{-(x-B)^2}{C^2}}$ | ICU Patients | ICU Patients + Deaths | H Patients + Deaths |
| --- | --- | --- | --- |
| $A$ | $4095 \pm 15$ | $20674 \pm 119$ | $48387 \pm 239$ |
| $B$ | $32.11 \pm 0.15$ | $39.78 \pm 0.19$ | $35.74 \pm 0.21$ |
| $C$ | $18.01 \pm 0.17$ | $18.58 \pm 0.14$ | $18.27 \pm 0.20$ |

Note: ICU Patients are Patients in Intensive Care Units, while H Patients are composed by Hospitalized Patients with symptoms and Patients in Intensive Care Units.

Finally we show what happened after that the first curve reached the turning point (**Fig. 3**). The curve a begins the decrease that is not Gaussian, but it is perfectly described by a straight line (**Table 3**); on the contrary, the other two curves do not have a rapid decrease. Due to the fact that the number of deaths continues to be very high, the curve b increases (**Table 3**) in an almost linear way (the last two points are perhaps not correct for the reason explained in the introduction), while the *curve c* has a plateau and a very slow degrowth.

**Figure 3.**
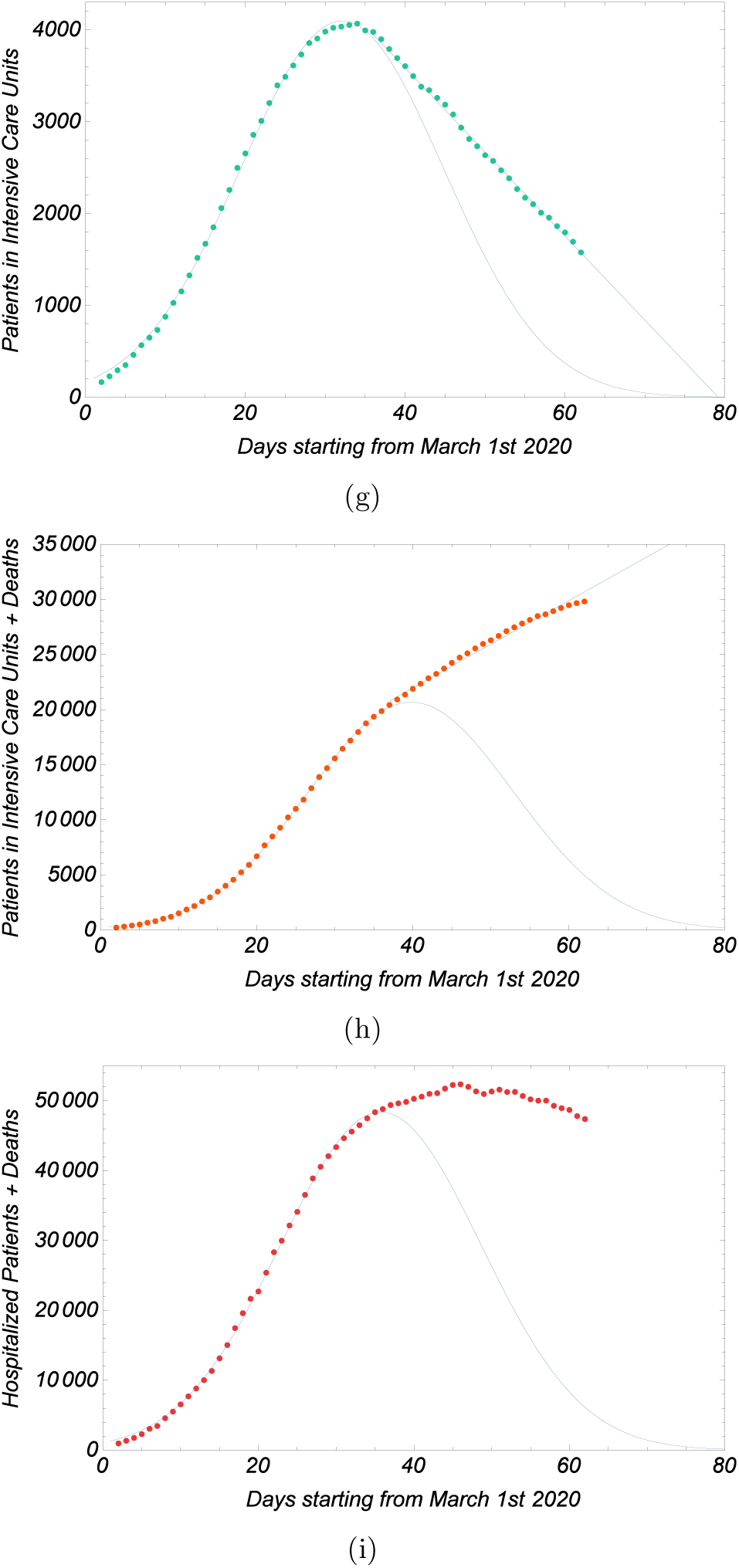
The Epidemic Spreading for all the examined interval of time: (a) Patients in Intensive Care Units, (b) Patients in Intensive Care Units and Deaths, (c) Hospitalized Patients (Hospitalized Patients with symptoms and Patients in Intensive Care Units) and Deaths.

**Table 3.** Parameters of least squares fits after the turning point of the Epidemic

| $y(x) = mx + b$ | ICU Patients | ICU Patients + Deaths |
| --- | --- | --- |
| $m$ | $-91.5 \pm 0.7$ | $396.18 \pm 8.98$ |
| $b$ | $7245 \pm 36$ | $6119 \pm 442$ |

Note: ICU Patients are Patients in Intensive Care Units.

## 3 Conclusions

We have analyzed some data concerning the pandemic of COVID-19 in Italy provided by the Italian Civil Protection. In particular, we have followed the evolution of three important parameters, such as ICU patients (*curve a*), ICU patients plus deaths (*curve b*), and the sum of hospitalized patients and the deceased (*curve c*). We have observed a similar Gaussian behavior in the growth phase of the statistical curves, but a totally different behavior in the phase after the turning point. We can summarize the results of our analysis in the following points.

1. Among the three examined curves, only the first begins (as expected) a rapid decrease that coincides also with the behavior of the number of infected people (the Italian Civil Protection announced that the epidemic has achieved its peak on March 31st), so it can be considered as a good thermometer of the trend of the virus in Italy and can be suggested as a useful tool to understand the evolution of epidemic also for other countries.
2. In particular, the extrapolation of data for curve a allows to predict the day of zero ICU in Italy, because the straight line of decrease finishes on day 80, that is on May 19th. Of course, also this prediction will probably prove wrong, because the end of the first phase of lockdown in Italy on May 4th may change the present trend of the curve.
3. Another important aspect of the first curve is its turning point, because it fixes also the change of the trend of the other two curves.
4. Surprisingly, after the turning point, the direction taken by the three curves is very different: the first decreases, the second increases and the third stays almost constant and begins a slow degrowth. This behavior can be explained in the light of the too high number of deaths, an anomaly if compared with all the other indicators at the end of April.
5. Even if the *curves b* and *c* have not respected the predicted trend and cannot be used to predict the future evolution, they should not be neglected. In fact, it is very difficult to imagine a new phase for the epidemic of COVID-19 in Italy without a decrease of the *curve b* and *c* that is if the number of deaths everyday is not at least less than the number of people that get out from the Hospital alive. On the other side, those curves could be used also with a predictive power in a different context where the mortality rate is not as high as in Italy.

## Data Availability

database provided by the Italian Civil Protection

## Notes

### Competing Interest Statement

The authors have declared no competing interest.

### Funding Statement

University of Benevento

